# Integrative Analysis of the Blood Proteome by Mendelian Randomization Reveals Regulatory Networks in Calcific Aortic Valve Disease

**DOI:** 10.1101/2023.07.26.23293222

**Authors:** Mewen Briend, Louis-Hippolyte Minvielle Moncla, Valentine Duclos, Samuel Mathieu, Anne Rufiange, Sébastien Thériault, Benoit Arsenault, Yohan Bossé, Patrick Mathieu

## Abstract

**Background:** Calcific aortic valve disease (CAVD) is a disorder characterized by fibrocalcific remodeling of the aortic valve (AV). The blood molecular phenome involved in CAVD is presently unknown.

**Methods:** We carried out a proteome-wide two-sample Mendelian randomization (MR) study to identify circulating molecules causally associated with CAVD. We queried as the exposition a large cohort of 35,559 subjects in whom 4,719 blood proteins were measured. For the outcome, we leveraged a recent GWAS for CAVD including 13,765 cases and 640,102 controls. Single-cell RNA-seq was analyzed to highlight potential pathways affected by the blood proteome.

**Results:** In MR, we identified 49 blood proteins robustly associated with the risk of CAVD. The blood proteins formed a network enriched in the immune response and ligand-receptor interactions. PCSK9, APOC3, ACE and IL6 were identified as actionable targets suitable for drug repurposing. Modulators of innate (IL6R, CNTFR, KIR2DL3-4) and adaptive (IL15RA, IGLL1, LILRA6) immune responses were associated with CAVD. Different regulators of platelets activity such as soluble GP1BA, COMP and VTN were also related to the risk of CAVD. Circulating modulators of the transforming growth factor-beta (TGF-beta) family such as ASPN, LEFTY2 and FSTL3 were associated with the risk of CAVD and their directional effects were consistent with the role of this pathway in the pathogenesis. Analysis of ligand-receptor interactions in the AV, which was inferred from single cell RNA-seq, provided further evidence that the IL6 and TGF-beta pathways are activated in CAVD.

**Conclusions:** We identified 49 blood proteins robustly and causally associated with CAVD, which were involved in the metabolism of lipids, immunity, regulation of blood pressure, platelet activation and modulation of growth factors activity. The present MR scan of the blood proteome provides a roadmap for follow-up studies and drug repurposing in CAVD.

**Clinical Perspective:** *What is new?:*

- The causal blood molecular phenome is presently unknown in CAVD; herein we investigated by Mendelian randomization the causal associations between the blood proteome and the risk of CAVD.
- In total, 49 blood proteins were found causally associated with the risk of CAVD and were involved in the metabolism of lipids, control of the immune response, regulation of blood pressure, platelet activity and the modulation of growth factors activity.
- Single cell RNA-seq analysis of calcific aortic valves revealed several ligand-receptor interactions potentially affected by the blood phenome.

*What are the clinical implications?:*

- There is no drug therapy available to treat CAVD.
- Analysis of the blood proteome by Mendelian randomisation showed that in-development, approved drugs or biologics targeting PCSK9, APOC3 and ACE could be repositioned and investigated in order to treat CAVD.

## Introduction

Calcific aortic valve disease (CAVD) is a disorder characterized by a fibrocalcific transformation of valve interstitial cells (VICs), which leads to the development of aortic valve stenosis^1, 2^. Mineralization and fibrotic processes of the aortic valve (AV) are promoted by the transforming growth factor beta (TGF-beta) pathway^3^. Different upstream factors involving lipids, inflammation and activation of platelets have also been shown to promote fibrocalcific transition of VICs and the development of CAVD in different pre-clinical models^4–7^. A vast repertoire of circulating proteins with diverse regulatory functions interact with proteins expressed by blood cells and peripheral tissues^8^. As such, circulating proteins provide a layer of control on several key cellular functions. The causal blood molecular phenome, which interacts with the AV and promotes CAVD, is presently unknown.

Mendelian randomization (MR) is a technique relying on the use of gene variants as instrumental variables in order to infer causal relationships between exposition and outcome variables^9^. As gene variants are randomly allocated in the population well before the development of the outcome, MR is not prone to reverse causality bias^10^. MR has been previously implemented in order to identify causal risk factors in CAVD. MR-based inference showed that low-density lipoprotein cholesterol (LDL-C), lipoprotein(a) [Lp(a)], obesity and blood pressure were causal risk factors in CAVD^11–13^. However, the circulating molecules that play a causal role in the development of CAVD are still unidentified. Herein, we undertook a proteome-wide MR study to identify the causal molecular blood phenome robustly associated with the risk of CAVD. Data were crossed with single-cell RNA sequencing (scRNA-seq) in CAVD to infer ligand-receptor interactions. We identified 49 blood proteins, some of which are actionable, involved in the metabolism of lipids, control of the immune response, regulation of blood pressure, platelet activity and modulation of growth factors activity.

## Methods

### Genome-wide association study for CAVD

Summary statistics for a GWAS meta-analysis including 13,765 cases in 640,102 European-ancestry participants from ten cohorts were downloaded ^11^. The study reported 11,591,806 variants. Genomic coordinates in GRCh37 were converted in GRCh38 by using LiftOver tools from UCSC.

### Proteome-wide Mendelian randomization

Summary statistics from deCODE genetics encompassing genetic association analyses performed for 4,719 blood proteins measured by an aptamer-based method (SomaScan) (4,907 aptamers) in 35,559 Icelanders were downloaded^14^. Two sample MR was performed by using a minimum of 3 *cis*-independent genetic variants as instrumental variables (P<5e-8) located within 250 kb upstream or downstream of the corresponding protein-encoding transcription start site. Independent variants were selected based on linkage disequilibrium (LD) (r^2^<0.1) by using PLINK1.9 and genotypes from European populations from the 1000 Genome project. The F-statistic was estimated by using the formula Beta^2^/SE^2 15^. We measured heterogeneity by using the Cochran’s Q test and pleiotropy with the Egger-intercept test. We implemented the inverse variance weighted MR and the weighted median MR. False discovery rate (FDR) and Bonferroni correction were calculated by using the R packages multtest with the Benjamini and Hochberg test. We performed MR analyses by using the Mendelian Randomization package.

### Enrichments

Enrichment analyses were performed by using data from InnateDB^16^ (innate immunity genes) and ligand-receptor pairs as reported by Shao et al^17^. Hypergeometric tests (python library scipy.stats) were performed by using all the protein coding genes as the background. Enrichment for Gene Ontology and pathways were performed with Enrichr. Enrichment results were visualized using the ggplot2 package.

### Network

We generated the network by using causally associated blood protein as seeds in the InnateDB curated human protein-protein interactions dataset. Data visualisation was performed with Cytoscape 3.9.1 alongside StringApp and Enhanced graphics addons. Metrics and algorithms including degree centrality, cliques and voterank, were determined with python library NetworkX. Module analysis was performed with the walktrap algorithm implemented in NetworkAnalyst.

### Single-cell data analysis

We downloaded single-cell sequencing data for the 4 samples corresponding to calcific aortic valve specimens from BioProject (PRJNA562645). Parallel-fastq-dump (v0.6.5) was used to extract FASTQ from SRR files. Cellranger (v7.0.0) was used to perform alignment in hg38, filtering, barcoding and UMI counting from FASTQ. Downstream analyses were performed with the Seurat library (4.0). Cells with less than 150 features, greater than 5 percent of mitochondrial genes or 15 percent of ribosomal genes were excluded. Cells were log-normalized with the scale factor 10,000, followed by a selection of 2,000 highly variable features with the vst method. Variables were scaled and centered to run a PCA dimensionality reduction, which was analyzed by using elbow plot feature and Jackstraw to select 20 dimensions and to perform 2D cell projection with clustering (Louvain algorithm, resolution 0.5) and tSNE representation. Markers for each cluster were obtained by using Wilcoxon Rank Sum. Differentially expressed genes between clusters were used to manually annotate each cluster.

### Single-cell ligand-receptor communication

Inferred ligand-receptor interaction in single-cells was performed by using CellChat (1.6.1). 1,939 validated molecular interactions from CellChat database were used. Communication probability was measured with 5% truncated mean and data were represented in circular visualization.

### Identification of actionable targets

We interrogated DGIdb, an extensive repository of drug-gene pairs collated through different resources, to assess actionable targets from the causal blood proteins. We considered approved and in-development drugs. A target was deemed actionable if reported in DGIdb or in the literature and were in-development or approved drugs.

## Results

**Figure 1** presents a schematic of the study design.

**Figure 1:**
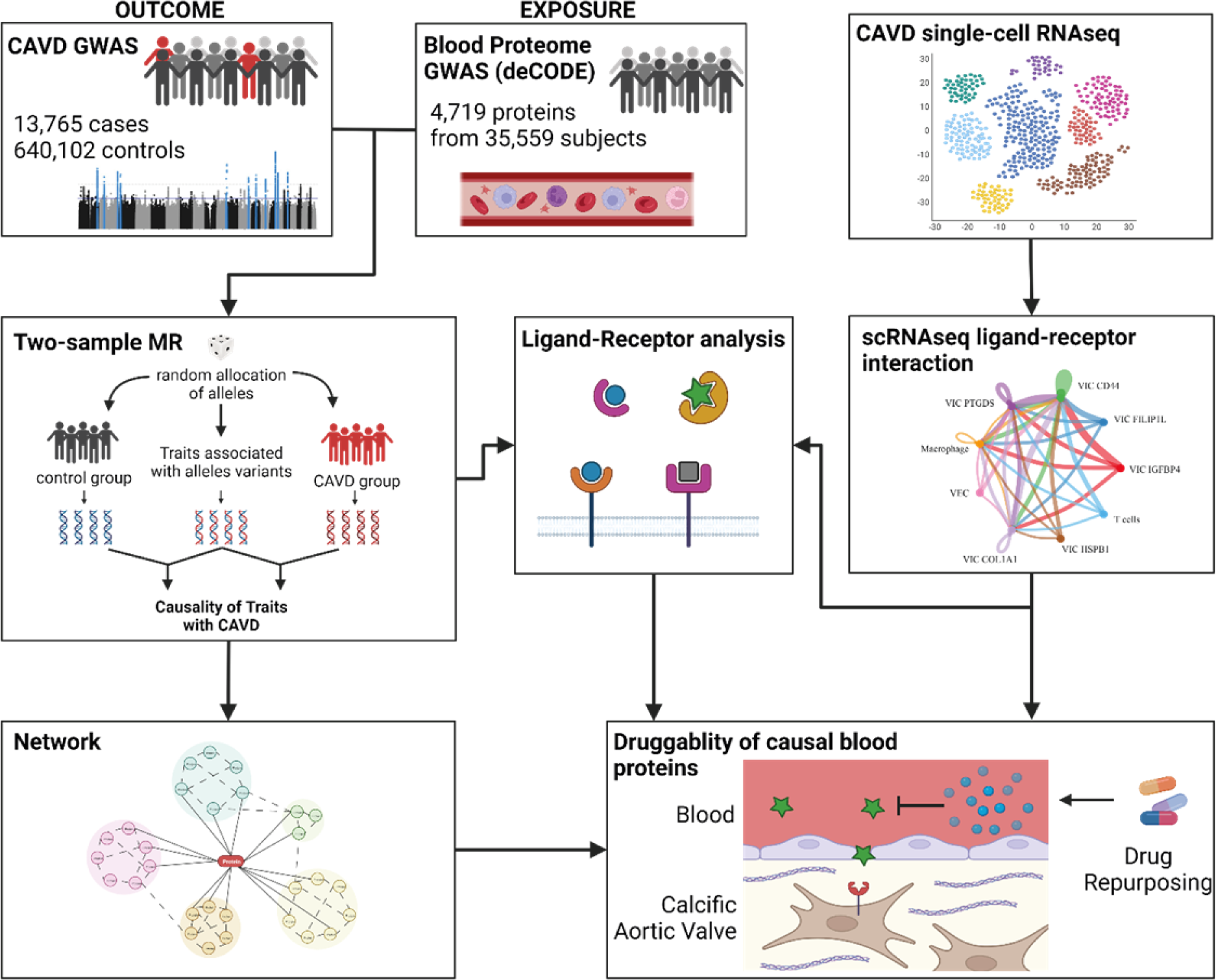
Study plan. Schematic representation of the analytical pipeline. The left-hand panel represent the Mendelian randomization analyses using as exposition blood proteins and as outcomes CAVD GWAS. Causal blood proteins obtained were integrated into network and ligand-receptor analyses. The right-hand panel relies on the analysis of single-cell RNA-sequencing from four diseased aortic valves with inference of ligand-receptor interactions. Data were integrated for biological understanding and to identify potential drug targets.

### Proteome-wide Mendelian randomization

We carried out a MR analysis of the blood proteome in CAVD. We leveraged a recent GWAS for CAVD totalling 13,765 cases and 640,102 controls of European ancestry^11^. For the exposition, we queried a large cohort of 35,559 subjects of European ancestry in whom protein quantitative trait loci (pQTL) were obtained for 4,719 blood proteins measured by an aptamer-based method^14^. We performed inverse variance weighted (IVW) MR by selecting independent *cis* instrumental variables (P<5E-08) located within ± 250kb from the transcription start site of the gene encoding the protein of interest (Methods). Analysis was performed for 1,313 different blood proteins for which we had enough instrumental variables (≥3) to carry out multi-instrument MR (median instrumental variables: 31). The F statistics was greater than 29 for all the instrumental variables, which is well above the minimal threshold of 10 recommended for MR analysis^15^. After Bonferroni correction for multiple testing, 19 blood proteins were associated with CAVD **(Figure 2A).** By using a false discovery rate (FDR) < 5% for multiple testing correction, 66 blood proteins were associated with CAVD **(Supplementary Table 1) (Figure 2A)**. To evaluate the risk of heterogeneity we implemented the Cochran’s Q test (IVW and Egger tests of heterogeneity). From the blood proteins with FDR <5%, 60 did not show pleiotropy (Cochran’s Q P > 0.05) **(Supplementary Table 1)**. For additional sensitivity measure we implemented the weighted median MR, which is robust to invalid instruments^18^. Among the blood protein candidates (FDR_IVW_<5%, Cochran’s Q P > 0.05), 49 were at least nominally significant (P_weighted median_<0.05) in weighted median MR and were considered as causal candidates for downstream analyses **(Figure 2B)**. The Egger intercept test showed absence of pleiotropy for 45 blood candidates (P_intercept_ > 0.05), whereas 4 blood proteins showed modest pleiotropy **(Supplementary Table 1)**. The strongest association (lowest P_IVW_) with CAVD was soluble IL6R (per 1SD, OR: 0.94, 95%CI:0.92-0.96, P=7.99E-10). The largest positive and negative effect size proteins were circulating BDNF (OR: 1.79, 95%CI:1.36-2.36, P=4.03E-05) and FSTL3 (follistatin like 3) (OR: 0.65, 95%CI:0.51-0.84, P=7.90E-04), respectively a neural growth factor and an inhibitor of activin A signaling^19, 20^.

**Figure 2:**
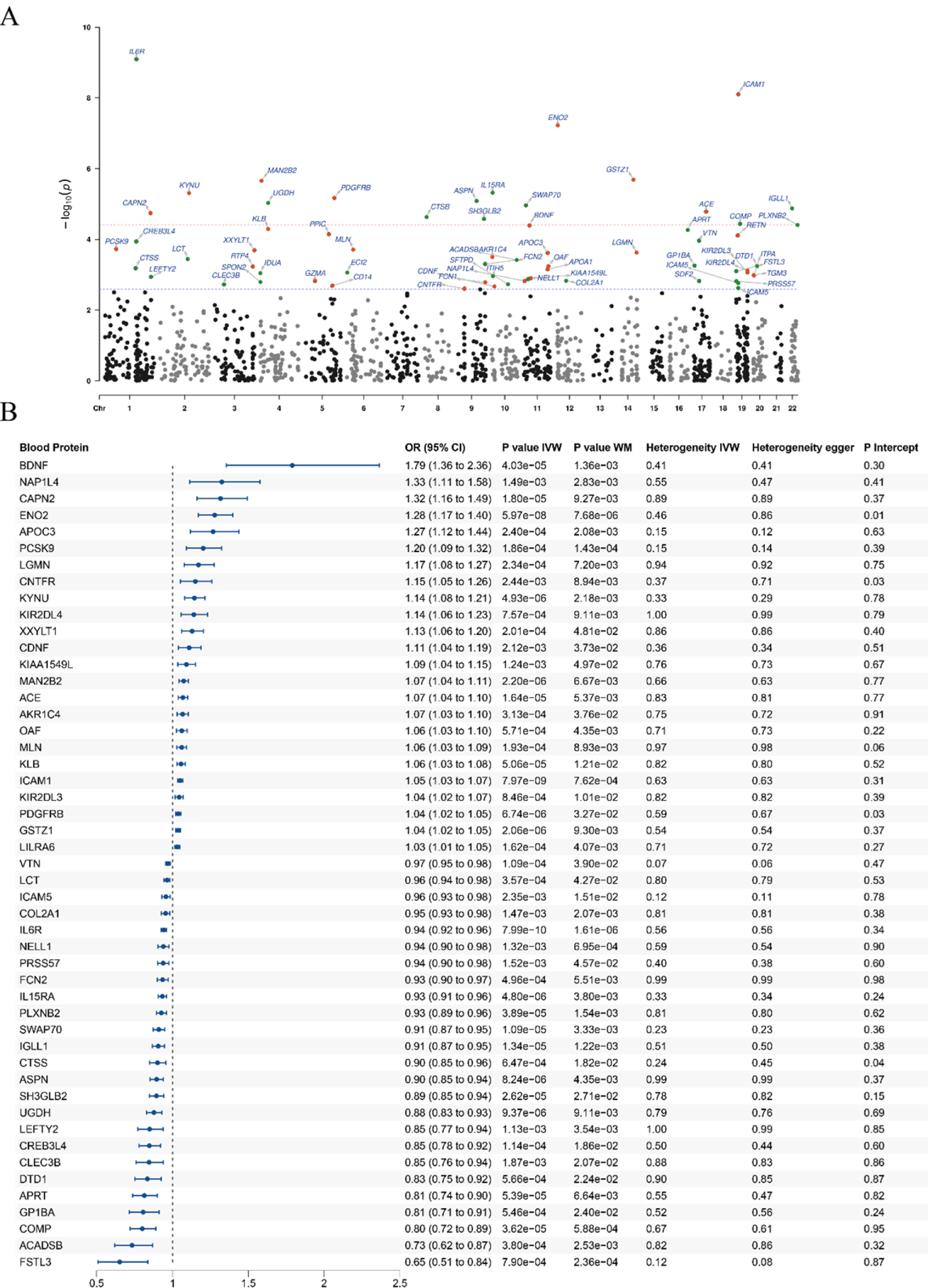
Blood proteins causally associated with CAVD. A) Manhattan plot showing 66 blood proteins significantly associated with CAVD by MR. Green and red dots illustrate blood proteins negatively and positively associated respectively. Blue and red dashed lines represent the P-value adjusted with FDR 5% and Bonferroni respectively. B) Forest plot illustrating significant MR association between 49 blood proteins and CAVD after heterogeneity–filtering. OR: odds ratio; 95%CI: confidence intervals at 95 %; IVW: inverse variance weighted; WM: weighted median.

### Evidence-based network for CAVD-associated blood proteins

We hypothesized that the blood proteome associated with CAVD in MR was enriched for the immune response. By using the immune dataset from InnateDB as the reference, we found that the CAVD-blood proteome was highly enriched in immune response associated molecules (ACE, CTSS, GP1BA, ICAM1, IL6R, IL15RA, IGLL1, KIR2DL3, KIR2DL4, LILRA6, PDGFRB) (fold-enrichment: 5.35, P=4.44E-06, hypergeometric test). In order to assess the function of the blood proteome, we extracted a network from the InnateDB dataset. The blood protein-protein interaction network for CAVD included 544 nodes (35 causal blood proteins) and 596 edges (connections) **(Figure 3A)**. We assessed the pathway enrichment of blood CAVD-network by using the Reactome dataset. This analysis showed that the network was enriched in signaling by receptor tyrosine kinases (P=1.56E-34), immune system (P=1.24E-30) and cytokine signaling in immune system (P=2.65E-30) **(Figure 3B)**. We implemented a community-searching algorithm for the blood CAVD-network in order to identify modules with specific functions (Methods). From the network, we identified 24 significant modules (P<0.05) **(Figure 3A, Supplementary Table 2)**. **Figure 3C** represents the gene ontology (GO) enrichment for each module. For instance, module 16 is enriched in regulation of interleukin-6 production (GO:0032675) (P=3.53E-09) and module 13 which includes the CAVD-associated blood protein PLXNB2 (OR: 0.93, 95%CI:0.89-0.96, P=3.89E-05) is enriched in semaphorin-plexin signaling pathway (GO:0071526) (P=2.20E-06). Among the other modules involved in immune regulation, module 18 includes CAVD-associated blood protein IL15RA (OR: 0.93, 95%CI:0.91-0.96, P=4.80E-06), which is enriched in myeloid leukocyte differentiation (GO:0002573) (P=3.55E-06) and module 20 includes CAVD-associated protein KIR2DL4 (OR: 1.14, 95%CI:1.06-1.23, P=7.57E-04), which is enriched in positive regulation of natural killer cell cytokine production (GO:0002729) (P=3.78E-06). Next, we implemented the voterank algorithm, an iterative process that prioritizes influential nodes (proteins)^21^. From the 544 nodes, voterank prioritized 39 blood proteins of which 28 were CAVD-associated blood proteins in MR **(Supplementary Table 3)** . These proteins were enriched in ciliary neurotrophic factor-mediated signaling pathway (GO:0070120) (P=1.88E-05), positive regulation of chemotaxis (GO:0050921) (P=3.35E-05), regulation of immune response (GO:0050776) (P=1.07E-04) and positive regulation of smooth muscle cell migration (GO:0014911) (P=2.53E-04) **(Suppl. Figure 1)**. Among the voterank blood proteins, 9 (KYNU, SWAP70, VTN, ENO2, SH3GLB2, UGDH, PDGFRB, CAPN2, NAP1L4) were within the top 10 proteins with the highest degree (highest number of communications) in the network. For instance, KYNU (kynureninase) (OR: 1.14, 95%CI:1.08-1.21, P=4.93E-06) is a voterank and highly connected protein involved in the metabolism of tryptophan and generates anthranilic acid (AA) and 3-hydroxy anthranilic acid (3-HAA)^22^.

**Figure 3:**
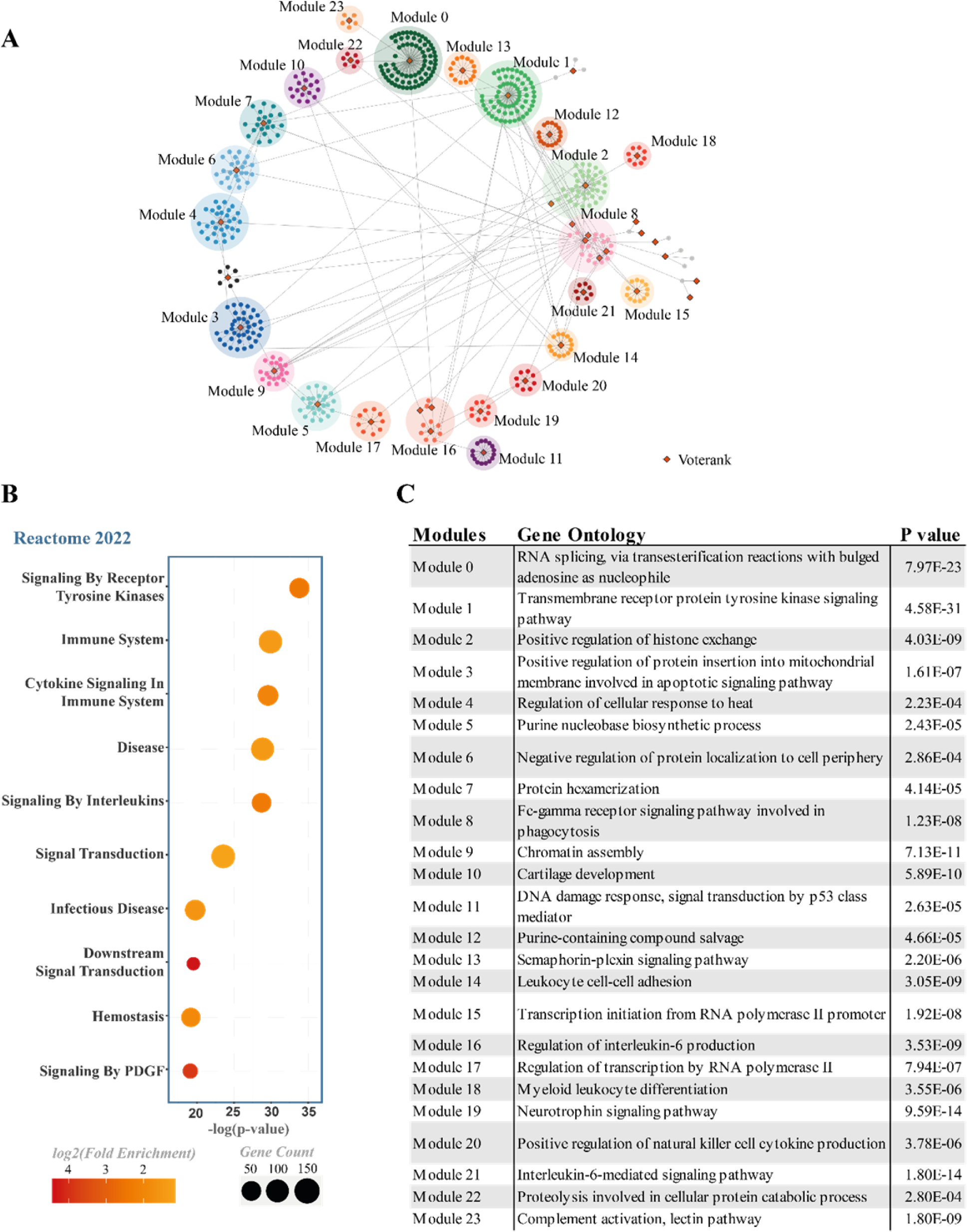
Protein-protein interaction network analysis. A) Causal blood proteins were used as seeds to generate a protein-protein network from InnateDB. Influential spreaders were identified by using the voterank algorithm and are represented by orange diamonds. Modules were identified using the walktrap algorithm and are represented by colored circles. B) Reactome enrichment analysis of the full network. Color represents the log2 fold-enrichment; circle size represents the number of nodes overlapping the terms and the x-axis represents the P-value for enrichment (Fisher’s exact test). C) Gene Ontology biological process enrichment analysis of all network modules.

### Ligand-receptor interactions

The blood proteome is involved in cell communication as it transports ligands and receptors. Soluble receptors play an important role in modulating the biological response^8^. Soluble receptors may dampen signaling by acting as decoy molecules for the natural ligands, whereas in some conditions they may promote signaling by being recruited along with their ligands to the cell membrane (trans-signaling)^23^. We assessed if the CAVD-associated blood proteome was enriched in ligands and receptors by interrogating a comprehensive compendium of ligand-receptor interactions^17^. We found that the CAVD-associated blood proteome was highly enriched in ligands and receptors (fold-enrichment: 5.20, P=6.26E-11, hypergeometric test). In total, CAVD-associated blood proteins in MR were involved in 90 potential ligand-receptor interactions. **Figure 4A-B** presents the inferred interaction pairs by using the CAVD-associated blood ligands and receptors identified in MR. CAVD-associated blood ligands such as COL2A1, COMP and VTN (vitronectin) are extracellular matrix-interacting proteins. VTN (OR: 0.97, 95%CI:0.95-0.98, P=1.09E-04), a protein involved in angiogenesis, tissue repair and thrombus formation^24^, is predicted to interact with several receptors including different integrins, TNFRSF11B (OPG), PLAUR and CD47. COMP (cartilage oligomeric matrix protein) (OR: 0.80, 95%CI:0.72-0.89, P=3.62E-05), a natural inhibitor of thrombin^25^, is predicted to interact with ITGB1 and ITGB3. FCN2 (ficolin 2/L-ficolin) (OR: 0.93 95%CI:0.90-0.97, P=4.96E-04), a member of the soluble defense collagens, interacts with LRP1, a receptor involved in phagocytosis^26^. LEFTY2 is a circulating ligand negatively associated with the risk of CAVD (OR: 0.85, 95%CI:0.77-0.94, P=1.13E-03) interacts with type I (ACVR1B/ACTR-IB) and II activin receptors (ACVR2A/ACTR-IIA, ACVR2B/ACTR-IIB) whereby through competitive binding it prevents NODAL and TGB-beta signaling^27, 28^. Two ligands, APOC3 (OR: 1.27, 95%CI:1.12-1.44, P=2.40E-04) and PCSK9 (OR: 1.20, 95%CI:1.09-1.32, P=1.86E-04) interact with LDLR (low-density lipoprotein receptor) and are involved in the metabolism of triglyceride-rich lipoproteins and LDL-C, respectively^29, 30^.

**Figure 4:**
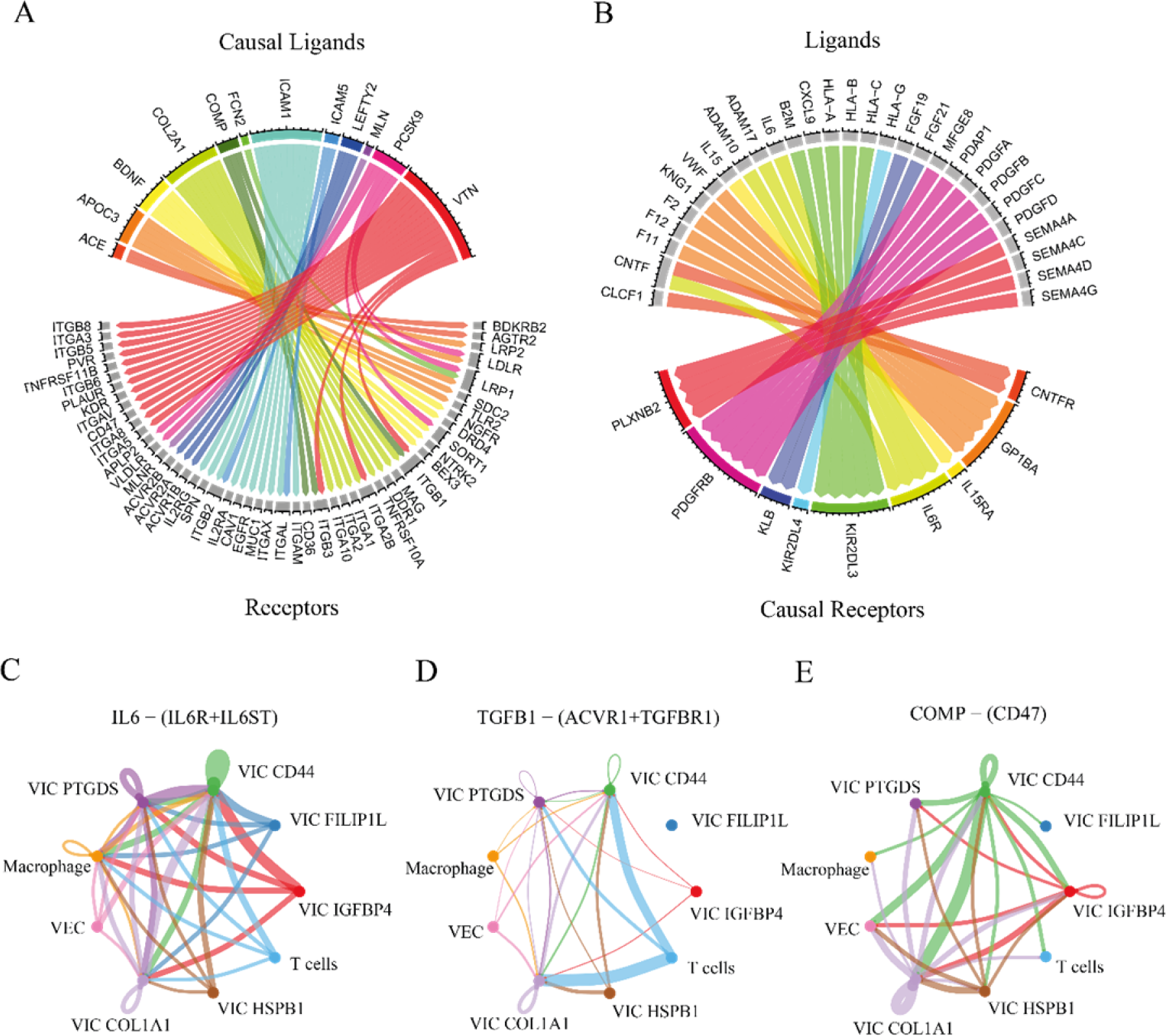
Ligand-receptor interactions identified from causal blood proteins and scRNAseq. A) Chord diagram illustrating causal ligand interactions with receptors and B) causal receptor interactions with ligands using a curated list of ligand-receptor interactions. C-E) Cell-cell communication in single-cell RNA-sequencing analysis inferred from CellChat showing interactions between IL6 and receptor (IL6R-IL6ST) (C), TGFB1 and receptor (ACVR1-TGFBR1) (D) and COMP with receptor CD47 (E); edge colors are consistent with the sources as sender; edges are proportional to the interaction strength.

Among the circulating receptors associated with the risk of CAVD in MR, soluble IL6R (OR: 0.94, 95%CI:0.92-0.96, P=7.99E-10), IL15RA (OR: 0.93, 95%CI:0.91-0.96, P=4.80E-06) and CNTRF (OR: 1.15, 95%CI:1.05-1.26, P=2.44E-03) are predicted to interact with IL6, IL15 and CNTF, respectively. In the blood, secreted IL6R and IL15RA form complexes with their target cytokines and act as decoys, which decrease signaling^31–33^. Secreted CNTFR has been shown to promote trans-signaling for CNTF, a cytokine of the IL6 family^34^. Soluble GP1BA (OR: 0.81, 95%CI:0.71-0.91, P=5.46E-04) was negatively associated with the risk of CAVD and is predicted to interact with several regulators of hemostasis including VWF (Von Willebrand factor). Soluble PLXNB2 (OR: 0.93, 95%CI:0.89-0.96, P=3.89E-05) is the receptor for the semaphoring group 4 (SEMA4). However, the role of soluble PLXNB2 is presently unknown.

We hypothesized that ligand and receptors expressed in the AV may be involved in the pathogenesis of CAVD and implicate cross-talk with the blood. We analyzed a publicly available scRNA-seq performed in 4 AVs from individuals with CAVD^35^. After quality check (Methods), 36,601 cells were clustered with the Louvain algorithm. We identified 9 clusters of which six were annotated to VIC subpopulations, one to valve endothelial cells (VEC), and two to immune cells (macrophages and T-cells) **(Suppl. Figure 2).** We next implemented the CellChat algorithm, which assesses cell-cell communication and provides a confidence score for each interaction based on gene expression level of ligands (sources) and receptors (receivers) retrieved from a curated compendium^36^. CellChat provided additional evidence that *IL6*, which is ubiquitously expressed by cells of the AV during CAVD, interacts with *IL6R* and its co-receptor *IL6ST* expressed by several subpopulations of VICs (*PTGDS*^+^, *CD44*^+^, *COL1A1*^+,^ *IGFB4*^+^) and T cells **(Figure 4C)**. Several interactions between *TGFB1* and *ACVR1*-*TGFBR1* were also predicted **(Figure 4D)**. T cells were found to be a major source of *TGFB1*, whereas VICs *CD44*^+^ and VICs *COL1A1*^+^ were predicted to be main receivers **(Figure 4D)**. COMP is a blood protein negatively associated with CAVD and according to CellChat *COMP* is also expressed in AVs during CAVD and is inferred to interact with integrins and *CD47* **(Figure 4E)**. In the AV, VICs *CD44*^+^ are the main source of *COMP*, which is predicted to interact with different subpopulations of VICs, valve endothelial cells and immune cells **(Figure 4E)**.

### Identification of actionable targets

We interrogated the Drug Gene Interaction Database, a curated resource collating the gene products that are interacting with drugs. By using the CAVD-associated blood proteins, we identified 18 different blood proteins representing 232 predicted drug-gene pair interactions **(Suppl. Table 4)**. Among the CAVD-associated blood proteins, ACE (OR: 1.07, 95%CI:1.04-1.10, P=1.64E-05), PCSK9 (OR: 1.20, 95%CI:1.09-1.32, P=1.86E-04), and IL6R (OR: 0.94, 95%CI:0.92-0.96, P=7.99E-10) are actionable targets for which approved drugs or antibodies could be repurposed for the treatment of CAVD. In the blood, soluble IL6R is considered as a natural inhibitor of IL6^31, 32^. Therefore, inhibition of IL6 signaling with blocking antibodies could be repurposed for CAVD. Circulating angiotensin converting enzyme (ACE) (OR: 1.07, 95%CI:1.04-1.10, P=1.64E-05) was positively associated with the risk of CAVD and is a target of several FDA approved ACE inhibitors, which could be investigated for drug repurposing. We found in MR a robust association between blood PCSK9 (OR: 1.20, 95%CI:1.09-1.32, P=1.86E-04) and CAVD. This finding is supported by previous MR analysis indicating that LDL-C and Lp(a) are likely causal in CAVD^12, 13^. APOC3 (OR: 1.27, 95%CI:1.12-1.44, P=2.40E-04), a regulator of blood triglyceride level^37^, was positively associated with the risk of CAVD and is a target of under-development oligo-antisense for the treatment of hypertriglyceridemia^38^. KIR2DL3 (OR: 1.04, 95%CI:1.02-1.07, P=8.46E-04) and KIR2DL4 (OR: 1.14, 95%CI:1.06-1.23, P=7.57E-04), which are targets of lilirumab, an antibody under development for indication in cancer^39^, were positively associated with CAVD and warrant further exploration in pre-clinical models. Manual curation for each blood causal candidate protein indicated that under development inhibitors for CNTRF (OR: 1.15, 95%CI:1.05-1.26, P=2.44E-03), a member of the IL6 family of cytokines, should be investigated for CAVD^40^. As soluble IL15RA (OR: 0.93, 95%CI:0.91-0.96, P=4.80E-06), a negative regulator of IL15 signaling, was associated with CAVD, inhibitors for IL15, which are in development for celiac disease, should be investigated in CAVD^41^.

## Discussion

In this work, by using a proteome-wide scan and MR we showed that 49 blood proteins were robustly associated with the risk of CAVD. Integration of data into networks showed enrichments in the immune response and ligand-receptor interactions. Blood proteins involved in the metabolism of lipoproteins, immunity, regulation of blood pressure, platelet activation and modulation of growth factors were causally associated with CAVD **(Figure 5)**. Integrative analyses showed that ligand-receptor interactions in the AV involving the IL6 and TGF-beta pathways are modulated by circulating proteins associated with the risk of CAVD. We identified drug targets suitable for repurposing and also druggable candidates, which warrant further investigation in order to develop novel therapies for CAVD.

**Figure 5:**
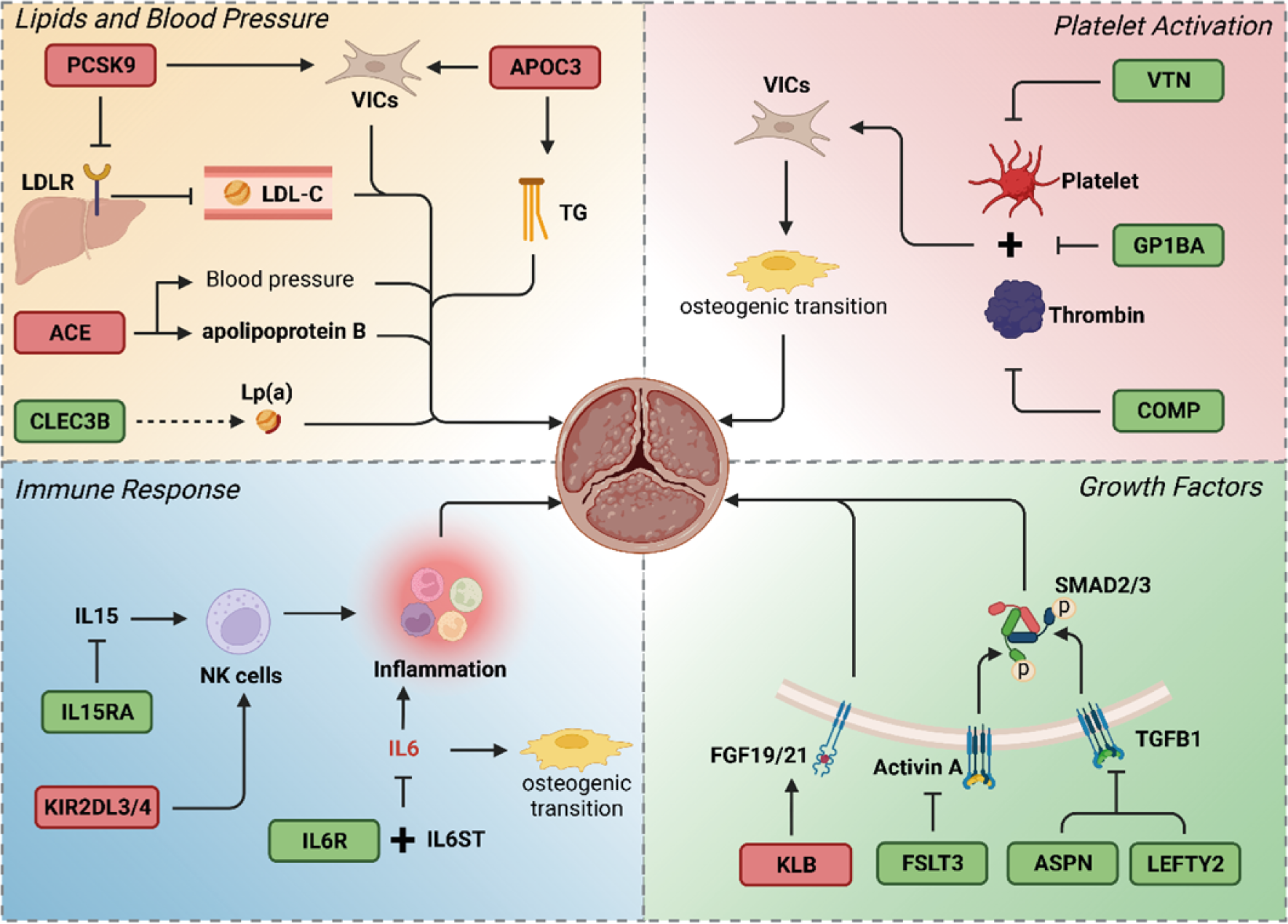
Integrative analysis and biological interpretation of results. Blood proteins in green and red rectangles are negatively and positively associated with CAVD respectively. The graph is divided into four parts, each representing a biological function. The first panel at the top-left shows the implication of lipids and blood pressure in CAVD. PCSK9 increases the *in vitro* calcification of VICs in cell culture^43^ and the LDL-cholesterol plasma levels by internalizing the LDL receptor^42^. APOC3 also promotes the calcification of VICs in culture and is involved in the TG plasma level^44^. CLEC3B interacts with apolipoprotein(a), which is involved in the development of CAVD^45^. The functional role of CLEC3B on apolipoprotein(a) remains unknown. ACE is an important regulator of the blood pressure, which is a major risk factor of CAVD^48^. ACE is transported by apolipoprotein B in AV^50^. The bottom-left panel represents the involvement of inflammation. IL6R forms a trimer with IL6ST and IL6 that reduces the signaling by IL6, a cytokine involved in the osteogenic transition of VICS and the mineralization of cell cultures^5^. Soluble IL15RA acts as a decoy for IL15, a cytokine implicated in the expansion and maintenance of cytotoxic T cells and natural killer cells^46^. KIR2DL3-4 modulate the activity of natural killer cells^47^, however their soluble role is unknown. The top-right panel represents the implication of platelet activation, a pathway known to promote osteogenic transition of VICs^7^. Soluble GP1BA prevents the adhesion between platelets and thrombin^53^. In the blood plasma, VTN reduces the aggregation of platelets^24^ and COMP inhibits thrombin^25^. The bottom-right panel shows the implication of growth factors in CAVD. KLB increases the activity of FGF19 and FGF21 (a fibroblast growth factor overexpressed in blood of CAVD patients) at their receptor^56^. FSTL3 is an inhibitor of activin A^55^. ASPN and LEFTY2 regulate TGFB1^28, 54^, an activator of SMAD2/3, supporting the implication of the super family of TGF beta in CAVD.

### Blood proteins involved in the regulation of lipids

We underlined that two key circulating regulators of lipid metabolism, PCSK9 (OR: 1.20, 95%CI:1.09-1.32, P=1.86E-04) and APOC3 (OR: 1.26, 95%CI:1.12-1.44, P=2.40E-04), were positively associated with the risk of CAVD. PCSK9 promotes the internalization of the LDL receptor (LDLR) and thus increases the blood LDL-C^42^. The present findings are in accordance with previous MR investigations, which showed that LDL-C was associated with the risk of CAVD^12, 13^. PCSK9 is also expressed in the aortic valve and promotes *in vitro* the mineralization of VIC cultures^43^. APOC3 is an important regulator of triglycerides^37^. The positive association between genetically predicted blood level of APOC3 with CAVD suggests that triglycerides are likely involved in the development of CAVD. Also, recent work suggests that APOC3 promotes the mineralization of VIC cultures by a pathway involving inflammation and mitochondrial dysfunction^44^. Among the CAVD-associated blood proteins, CLEC3B (tetranectin) (OR: 0.85, 95%CI:0.76-0.94, P=1.87E-03) interacts with apolipoprotein(a), a major protein of Lp(a), which is involved in the development of CAVD^45^. The functional role of CLEC3B on apolipoprotein(a) is not known. Further work is needed to elucidate the role of CLEC3B in CAVD.

### Immune response in CAVD: modulation by blood proteins

The blood proteins associated with CAVD were enriched in molecules involved in the immune response. Proteins involved in both the innate and adaptive immune responses were associated with CAVD. Soluble IL6R (OR: 0.94, 95%CI:0.92-0.96, P=7.99E-10) was negatively associated with the risk of CAVD. Studies have shown that in circulation IL6R forms a trimer with its co-receptor IL6ST and IL6 whereby it decreases signaling by IL6 and reduces inflammation^31, 32^. *In vitro*, IL6 promotes the osteogenic transition of VICs and the mineralization of cell cultures^5^. The role of IL6 in CAVD is further supported by the analysis of ligand-receptor interactions in scRNA-seq, which inferred cell-cell communication based on IL6 production. Soluble IL15RA (OR: 0.93, 95%CI:0.90-0.96, P=4.80E-06) is acting as a decoy for IL15^33^, a cytokine that promotes expansion and maintenance of cytotoxic T cells and natural killer cells^46^. Thus, the findings from this work suggest that cytotoxic T and natural killer cells are possibly involved in the development of CAVD. In support for a role of natural killer cells in CAVD, we found that blood levels of KIR2DL3-4 were positively associated with the risk of CAVD. Though the role of soluble forms of KIR2DL3-4 is not known, these proteins are involved in HLA class I interactions and modulate the activity of natural killer cells^47^. Further work is needed to decipher the role of immune response and KIR2DL3-4 in CAVD.

### Blood pressure

The blood pressure has been highlighted in a previous MR study as being an important contributing risk factor to CAVD^48^. Herein, we identified that circulating level of ACE (angiotensin converting enzyme) (OR: 1.07, 95%CI: 1.04-1.10, P=1.64E-05), an important regulator of the blood pressure^49^, increased the risk of CAVD. Previous work highlighted that ACE was likely transported in the AV by apolipoprotein B containing lipoproteins^50^. In an observational study, surgically explanted AVs from patients with CAVD were less remodeled in subjects treated with angiotensin receptor blockers^51^. Moreover, AVs from patients under angiotensin receptor blockers had less inflammation and expression of IL6. In a retrospective study, the use of ACE inhibitors was associated with a lower rate of AV mineralization^52^.

### Blood soluble factors with a role on platelets

Soluble GP1BA (glycocalicin) (OR: 0.81, 95%CI:0.71-0.91, P=5.46E-04), a platelet-derived protein, was negatively associated with CAVD. Soluble GP1BA prevents the adhesion of platelets to thrombin^53^. These data suggest that prevention of platelet activation decreases the risk of CAVD. Among the different blood proteins, VTN (OR: 0.97, 95%CI:0.95-0.98, P=1.09E-04) and COMP (OR: 0.80, 95%CI:0.72-0.89, P=3.62E-05), which were negatively associated with the risk of CAVD, are regulators of platelet activity. In the plasma, VTN prevents the aggregation of platelets, whereas COMP inhibits thrombin^24, 25^. Analysis of human mineralized AVs revealed the presence of platelet aggregates on the surface of valves^7^. Functional assessment showed that activated platelets promoted an osteogenic program by VICs and accelerated the development of CAVD in a murine model^7^. Taken together, these findings support a causal role for the activation of platelet in CAVD.

### Growth factors and their modulation by blood proteins

Single-cell analysis of ligand-receptor communication suggested a T helper 2 (T_H2_) response whereby *TGFB1* expressed by T cells was inferred to interact with VICs *CD44*^+^ and VICs *COL1A1*^+^. Among the important soluble regulators, LEFTY2 (OR: 0.85, 95%CI:0.77-0.94, P=1.13E-03), ASPN (OR: 0.90, 95%CI:0.85-0.94, P=8.24E-06) and FSTL3 (OR: 0.65, 95%CI:0.51-0.84, P=7.90E-04) modulate the activity of the super family of TGF beta molecules. LEFTY2 and ASPN negatively regulate TGFB1, whereas FSTL3 inhibits activin A^28, 54, 55^. KLB (klotho beta) (OR: 1.06, 95%CI:1.03-1.08, P=5.05E-05) increases the activity of FGF19 and FGF21 to their receptors^56^. The blood level of FGF21 is elevated in patients with CAVD^57^. However, the role of this system is not established in CAVD and further investigation is needed. Among the other growth factors with unknown role in CAVD, BDNF (OR: 1.79, 95%CI:1.36-2.36, P=4.03E-05) and CDNF (OR: 1.11, 95%CI:1.04-1.19, P=2.12E-03) are neurotrophic factors positively associated with the risk of CAVD. A study showed that BDNF promoted the mineralization of cementoblast cultures^19^.

### Actionable targets in CAVD

From this MR scan of the blood proteome, we identified a number of actionable targets in CAVD. In-development and approved drugs or biologics targeting PCSK9, APOC3 and ACE are potential targets in CAVD and warrant further clinical investigation. Also, biologics such as tocilizumab, which targets IL6 signaling, could be repurposed and investigated as novel immune modulatory agents in the treatment of CAVD. Some of the blood proteins, such as KIR2DL3-4, CNTF and those in the IL15 pathways are targetable and need further investigation to determine the molecular and cellular processes whereby they are involved in CAVD. Moreover, functional follow-up studies in pathways involving platelets and modulation of growth factors activity could lead to the development of novel therapies.

### Limitations

Though MR is a powerful technique of causal inference, some associations may be driven by horizontal pleiotropy. However, we implemented several sensitivity analyses such as the Cochran’s Q test and the Egger intercept test to detect heterogeneity and pleiotropy, respectively. In addition, the weighted median MR, which is robust to invalid instrument, was performed to decrease the risk of pleiotropy.

### Conclusions

Proteome-wide MR identified blood molecules robustly associated with CAVD and emphasized the role of lipids, inflammation, blood pressure, platelets and growth factors in the development of this heart valve disease. Molecules with unknown or poorly characterized functions, especially in the context of CAVD, warrant follow-up functional investigations. Some actionable targets were identified for drug repurposing. Hence, the present MR scan of the blood proteome provides a roadmap for follow-up investigations in CAVD.

## Data Availability

all data is pubicly available

## Acknowledgments

None.

## Supplemental Material

Figure S1-2

Table S1-4

